# Fine-grained spatial data-driven ensemble modeling for predicting Sylvatic Yellow Fever environmental suitability in Brazil

**DOI:** 10.64898/2026.03.26.26349443

**Authors:** Douglas Adriano Augusto, Livia dos Santos Abdalla, Eduardo Krempser, Pedro Henrique de Oliveira Passos, Daniel Garkauskas Ramos, Alessandro Pecego Martins Romano, Marcia Chame

**Affiliations:** Oswaldo Cruz Foundation, Rio de Janeiro, 21041-361, RJ, Brazil; General Coordination of Arbovirus Surveillance of the Ministry of Health, Brasília, 70719-040, DF, Brazil; General Coordination of the Strategic Health Surveillance Information Center of the Ministry of Health, Brasília, 70719-040, DF, Brazil

**Keywords:** Ensemble modeling, Fine-grained georeferenced data, Multi-scale spatial modeling, Machine-learning, Environmental suitability, Model sensitivity, Sylvatic Yellow Fever

## Abstract

Sylvatic Yellow Fever (YF) is an infectious mosquito-borne disease with significant epidemiological relevance due to its widespread distribution and high lethality for human and non-human primates, particularly in tropical regions of the planet such as in Brazil. Identifying regions and periods of high environmental suitability for the occurrence of YF is essential for preventing or mitigating its burden, as it enables the efficient allocation of surveillance efforts, prevention, and implementation of control measures. Environmental modeling of YF occurrence has proven to be an effective approach toward this goal; however, its effectiveness strongly depends on the modeling framework’s capabilities as well as the spatial and temporal precision of all associated data. We propose a fine-scale geospatial modeling of YF environmental suitability that is based on a generative machine-learning ensemble method built on a large set of high-resolution environmental covariates. First, we take the spatiotemporal statistical description of the environment of each of the 545 YF cases from 2019–2024 up to 30 m/monthly resolution at three buffer scales: 100 m, 500 m, and 1000 m ratios. Then, we perform a feature selection and train hundreds of One-Class Support Vector Machine submodels to form a robust ensemble model, whose predictions are projected to a 1×1 km resolution grid of Brazil under several metrics, exceeding seven million ensemble evaluations. The predictions ranked the Southern Brazil region with the highest mean suitability for YF, with a level of 0.64; Southeast comes next with 0.46, followed closely by Central-West region (0.44), North (0.39), and finally Northeast (0.28). The model exhibited high uncertainty for the North region, indicating that data collection efforts are much needed in this region. As for the environmental covariates, a feature analysis pointed out that *Land use and cover* accounts for the largest influence in the model output.

## 1. Introduction

Yellow fever (YF) is an arboviral disease of significant public health concern, particularly in tropical and subtropical regions. In Brazil, the sylvatic cycle of YF, involving non-human primates (NHP) and autochthonous mosquitoes, poses a persistent threat, with spillover events leading to human infections. The complex dynamics of sylvatic YF transmission is intrinsically linked to variable ecological conditions, including climate, vegetation, and human-induced landscape changes, which collectively define areas of environmental suitability for its occurrence. Recent outbreaks in Brazil have underscored the urgent need for robust, large-scale predictive tools to identify and monitor regions at risk, facilitating timely public health interventions.

Although a safe and effective vaccine against YF has been available for decades, modeling the disease remains critically important for public health planning, outbreak prevention and mitigation, particularly in endemic countries like Brazil, aggravated by its continental dimension. Vaccine coverage is still low in some regions due to logistical challenges, socioeconomic and cultural barriers, and vaccine hesitancy (aggravated by antivaccination movements), especially in remote or underserved regions. Moreover, ongoing environmental changes—such as deforestation, urban expansion, and climate variability—are altering the spatial dynamics of yellow fever transmission by expanding the range of both sylvatic vectors and NHP hosts. These factors elevate the risk of spillover events and raise concerns about the reurbanization of yellow fever, given the continued presence of *Aedes aegypti* and *Aedes albopictus* in densely populated urban areas [1]. In addition, the global yellow fever vaccine stockpile may be insufficient to contain large or simultaneous outbreaks [2], making it essential to prioritize vaccination efforts based on reliable risk assessments [3].

Accurate prediction of YF environmental suitability necessitates the integration of diverse and detailed spatial information. Handling the processing requirements arising from the vast amount of data is by itself challenging. Traditional modeling approaches often struggle to capture the intricate, non-linear relationships between a multitude of environmental variables and disease occurrence, especially when operating across varying spatial scales. Furthermore, the inherent uncertainties in ecological and epidemiological data demand sophisticated analytical frameworks that can account for model variability and enhance predictive reliability.

This study addresses these challenges by employing a fine-grained spatial data-driven ensemble modeling approach. By leveraging high-resolution georeferenced data, we aim to develop robust predictive maps of YF environmental suitability across Brazil. Our methodology consists of a machine-learning pipeline integrated with ensemble modeling technique targeted to uncover complex patterns in high-dimensional datasets. Ensemble methods combine the predictions of multiple individual models, thereby reducing bias and variance, and ultimately improving overall predictive performance and robustness [4]. The application of multi-scale spatial modeling will allow us to capture relevant processes operating at different geographical resolutions, providing a more comprehensive understanding of the environmental determinants of the disease. Through this comprehensive framework, we seek to not only predict environmental suitability but also to investigate model sensitivity to various input features, offering valuable insights into the key drivers of YF occurrence. This research contributes to the ongoing efforts to enhance surveillance, prevention, and response strategies for YF in Brazil by providing a large-scale, high-resolution predictive tool.

### Related works

Although georeferenced data on yellow fever occurrence in Brazil have been around for a long time, they were scattered and unsystematic—mostly from very few Brazilian states as well as from specific research efforts. Only recently the Brazilian Ministry of Health officially started reporting country-wide YF cases along with their spatial coordinates [5], opening a pathway for accurate fine scale analysis. One of the main challenges of YF prevention in humans is the capacity of targeting vaccine doses to the population in susceptible regions; the more precise the categorization of *susceptible region*, the most efficient is the prevention. However, only georeferenced case data can lead to this ideal to the fullest extent, because it allows capturing the precise landscape effects on YFV circulation.

One of the first study of the kind to work with georeferenced case data [6] combined various data sources (online sources, literature, WHO reports) to assemble 751 points (plus 404 polygons) spread over 47 countries, during the period of 1970– 2016. Although this is an interesting strategy in the face of data scarcity, it is hard to assess the overall quality of data due to their heterogeneity and uncertainty about how they were originally obtained. Also, the choice of such a long period of YF reporting data (almost half a century) might have led to a model that is too generic to precisely capture recent dynamics of the disease, which might have hindered its forecasting accuracy. Moreover, the work deals exclusively with human cases and focuses on the *Aedes aegypti* vector for model training, which differs from the known (sylvatic) yellow fever vectors in Brazil, *Haemagogus* and *Sabethes* spp.

In a related study [7], the authors modeled environmental suitability for YF during the 2008–2009 epizootic in southern Brazil, combining data on the distribution of NHP (*Alouatta* spp.), the vector (*Haemagogus*), climatic, topographical, and vegetation variables. A key aspect of the work is the incorporation of species distribution models (SDM) of NHPs and vectors as biotic covariates of the YFV distribution model. Although the authors claim that these covariates add information, ultimately leading to model improvement, it may not be generally applicable since when the YF cases originate from infected NHPs (and vectors), their presence are already implicit in the data and therefore the SDM variables should effectively collapse to a near constant “suitable” value at all case locations.

An investigation of the factors behind the YFV transmission was carried out in [8], where the authors combined environmental, ecological, and entomological data to study its spread in a portion of the Atlantic Forest biome in Brazil. They used georeferenced data on human, NHP, and mosquitoes infections, but rather than building a model to predict a risk map for YF, their focus was on the understanding of the epidemiology of the disease. They found that most of the human infections are within a 11 km radius of an infected NHP and conclude that there are two main conditions that favor YFV circulation: regions characterized by extensive, continuous forest cover, and those composed of smaller, fragmented forest patches.

While our modeling methodology shares many aspects with the mentioned studies, such as the use of georeferenced report data to build the models, our work improves upon what is in the literature in several points: firstly, we work on a very fine resolution of environmental covariates, up to 30 m spatial and monthly temporal resolution, including a large set of 51 classes which derivates 918 total variables through statistical augmentation; we employ a multiscale analysis to capture varying levels of landscape effects, in an ensemble composed of hundreds of machine-learning models; and, finally, we tackle the whole territory of Brazil, which has continental dimensions, providing country wide predictions on a 1×1 km resolution grid, which still could be further refined if desired.

## 2. Materials and Methods

Our main goal is to build a robust high-resolution ensemble model of sylvatic yellow fever suitability in Brazil. Firstly, this requires the statistical environmental characterization of all data points, which is split into two categories: *positive points* (confirmed YF occurrence points) and *prediction points* (map points where predictions of YF suitability will be made). To cope with the acute computational demand resulting from the high-resolution data as well as enabling fast and reliable model updating for timely surveillance, we had to develop a suite of parallel processing tools specifically tailored for the task. The remaining of this section describes the source data we used and how the ensemble model was built.

### 2.1. Environmental layers

To capture the precise surrounding information of the positive points without incorporating excessive noise, and also to enable fine-grained spatial predictions, we chose to work with the highest spatial resolution we had at our disposal, which is 30 m. While some layers were available natively at 30 m resolution, we downscaled the remaining others to 30 m merely to make the processing uniform, especially when computing the statistical description—some statistics are sensitive to the resolution, such as the sample size (*count*).

We relied on five data sources for environmental characterization:

- *Rainfall*: Precipitation data was derived from the Climate Hazards Group InfraRed Precipitation with Station data (CHIRPS v3) [9]. It is a 0.05^°^ resolution (∼5 km, downscaled to 30 m), quasi-global rainfall dataset spanning 1981 to near-present, which combines high-resolution satellite estimates (including geostationary and polar-orbiting data) with daily station gauge data from a variety of sources. The temporal resolution is monthly.
- *Temperature*: The Climate Hazards InfraRed Temperature with Stations (CHIRTS) [10] dataset provides a resolution of 0.05^°^ (∼5 km, downscaled to 30 m) of monthly minimum and maximum temperature data by merging satellite-based infrared observations with ground station measurements. Developed by the Climate Hazards Center, CHIRTS offers a consistent and spatially detailed record of minimum and maximum temperatures from 1983 onwards. Again, the temporal resolution is monthly.
- *Land use and cover*: Provided by the MapBiomas project (Collection 9 [11]) a collaborative initiative involving NGOs, universities, research institutions, and technology companies, aimed at producing high-resolution (30 m, natively) annual land cover and land use maps for Brazil from satellite images, with historical data available from 1985 onwards. The temporal resolution is annual.
- *Altitude*: NASA Shuttle Radar Topography Mission (SRTM) Version 3 comprises a high-resolution (30 m) elevation data of Earth.
- *Climate normals*: WorldClim 2.1 is a global climate dataset that provides 1 km gridded climate variables, including monthly averages of temperature and precipitation, based on historical data from 1970 to 2000. The dataset was developed using weather station records interpolated with elevation and other covariates, and is widely used in ecological and environmental modeling due to its spatial detail and global coverage.

All layers are in TIFF format (please refer to Appendix A to visualize their mini maps). Except for MapBiomas, which is originally in Albers, all layers were reprojected from WGS84 (EPSG:4326) to Albers (ESRI:102033) in order to make processing automation uniform.

### 2.2. Positive yellow fever cases

We used a dataset of 568 confirmed cases of yellow fever virus in NHPs (*n* = 560) and humans (*n* = 8), spanning September 2019 to January 2025, which comprises all georeferenced cases officially available from the Brazilian Ministry of Health. We split up the dataset into training (*n* = 545) and validation (*n* = 23) sets such that all cases in 2025 were used as validation. Figures 1 and 2 show, respectively, their distribution and summary of the number of cases per state. Due to under-reporting, which stems from the limitations and discrepancies of regional surveillance systems, it is important to emphasize that the official cases represent just a portion of the actual virus circulation in the territory during the period. One can notice that the South Region of Brazil (states of Paraná-PR, Santa Catarina-SC, and Rio Grande do Sul-RS) accounts for the majority of the reported cases in the period of the study (434 of 568 cases, or 76%). This unbalance tends to bias the model towards the environmental characteristics of the southern regions of Brazil that had YF cases. We could have lessened this effect by subsampling YF cases in the South, but due to the relatively few number of training cases we opted for building the model over the whole set of cases. Despite that, the model behaved fairly robustly, being capable of generalizing to environments far beyond South’s characteristics (see Sections 3 and 4 for more detail). Regarding the distribution across biomes, 78% of the cases concentrate in the Atlantic Forest, followed by Cerrado (18%), Pampas Grasslands (3%), and Amazon Rainforest (1%).

**Figure 1:**
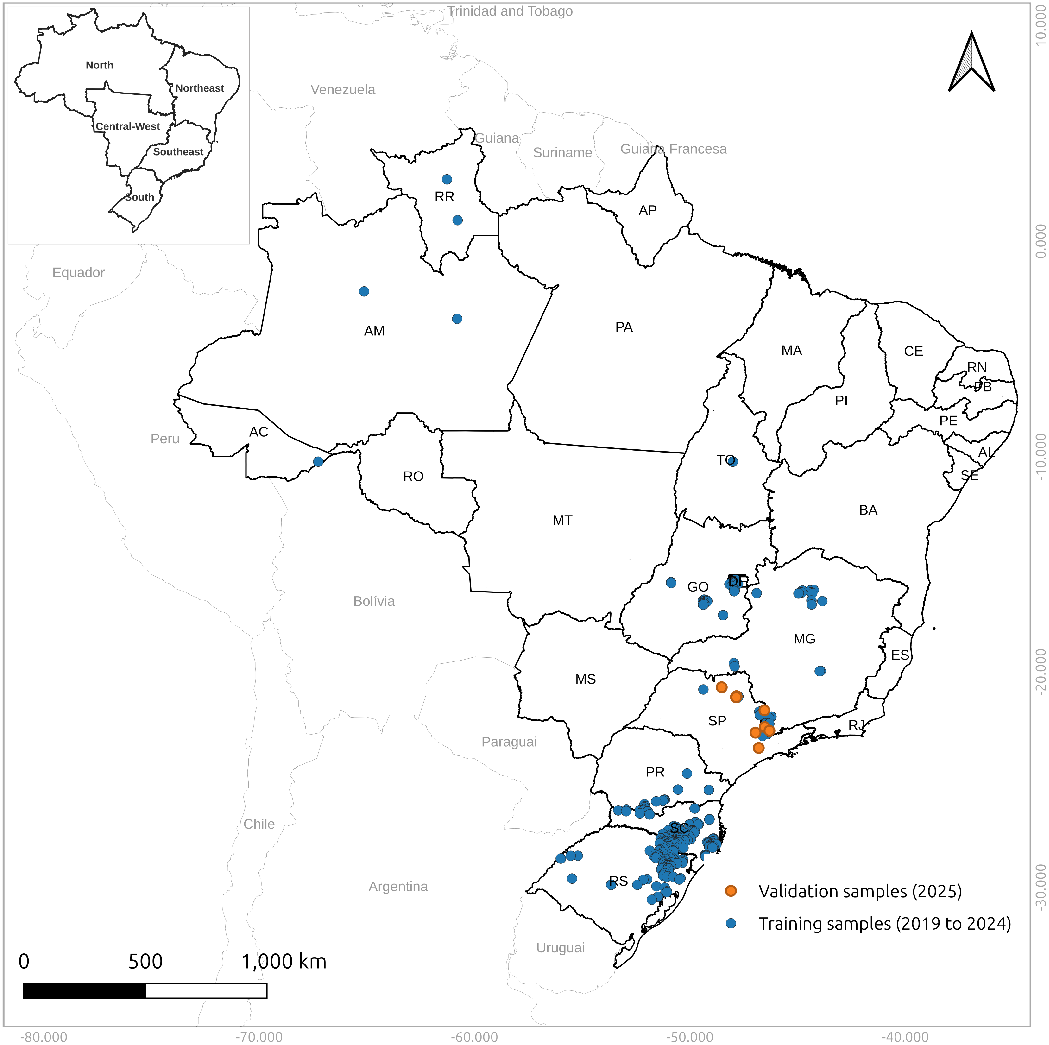
Distribution of the training and validation cases of YF.

**Figure 2:**
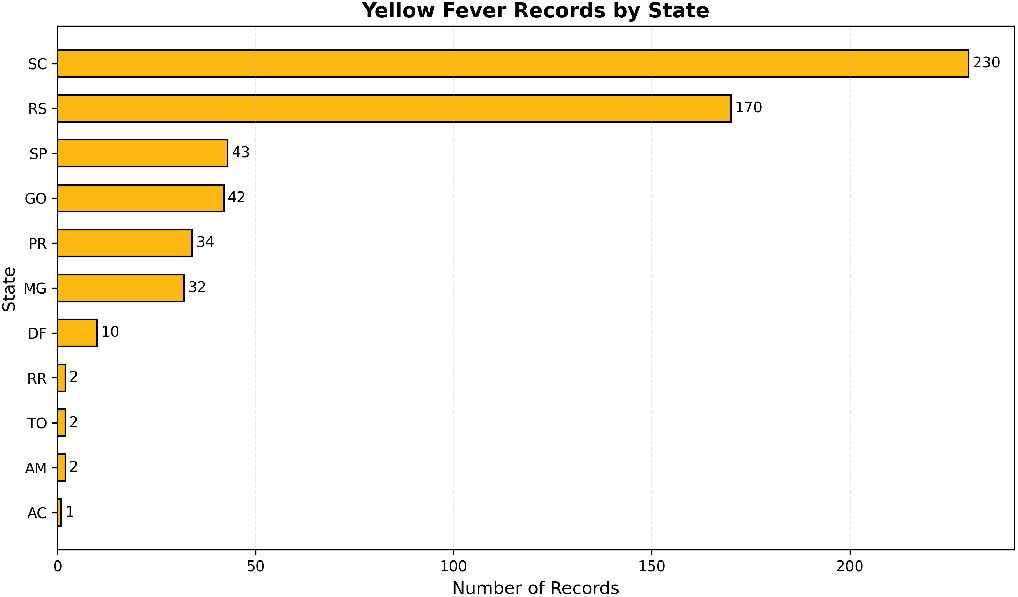
Summary of YF cases per Brazilian state (*n* = 568).

### 2.3. Steps summary

The whole modeling, depicted in Figure 3, can be summarized by the following steps:

1. Environmental characterization (zonal statistics)
  a. positive cases
  b. target grid (∼7.1 million points)
2. Merging
3. Filtering out features / feature selection
  a. Low-variability features
  b. Multi collinear features
4. Training & predictions
5. Analysis of impact of variables & knowledge extraction

**Figure 3:**
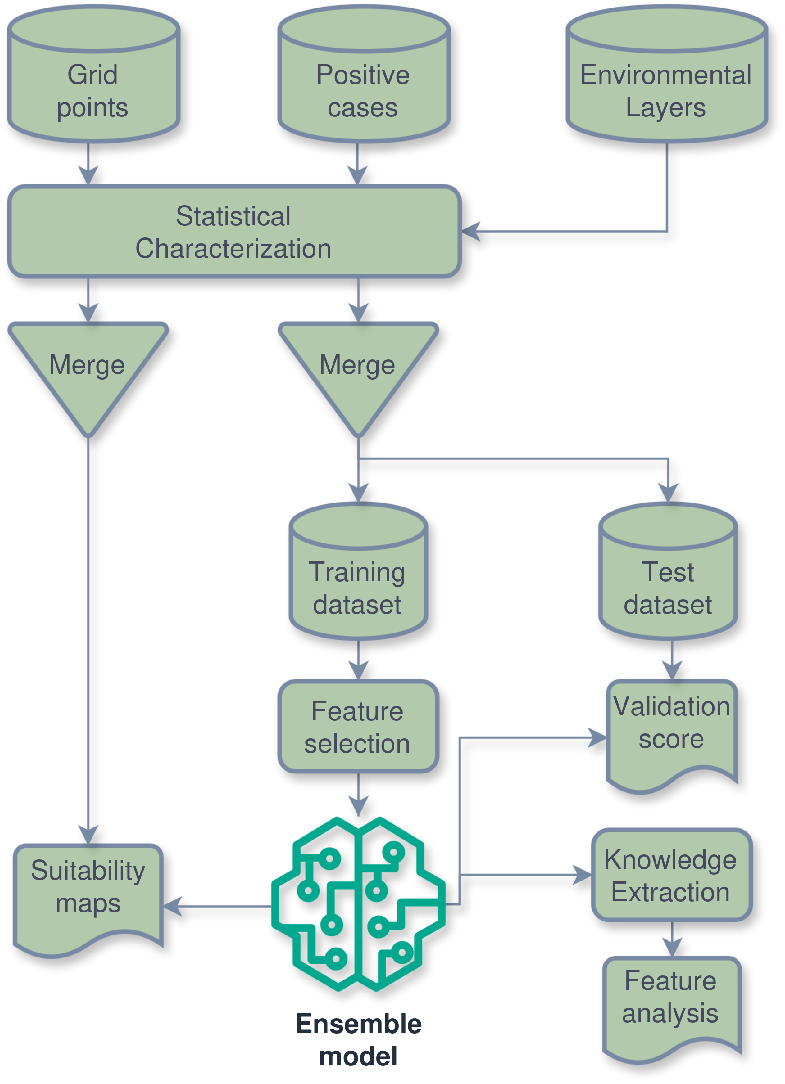
Flowchart of the modeling process.

### 2.4. Spatio-temporal statistical characterization

In the *characterization* step, we calculate 18 descriptive statistics within a buffer centered at the point of interest, which may be a positive (YF case) or grid point (for which predictions will be made). Nine of the eighteen descriptive statistics are normalized by the buffer area, allowing comparison across different buffer sizes. The statistics are [12]: *count, mean, minimum, maximum, percentile 25, percentile 50* (median), *percentile 75*, and *standard deviation*.

To embrace the concept of *landscape influence* as well as to account for data noise (such as GPS precision errors), we consider circular buffers around the points to calculate the zonal statistics. Moreover, to improve model robustness, we rely on a multiscale modeling consisting of three buffer size ratios: 100 m, 500 m, and 1000 m, which also take into account vector (mosquitoes) and host movements. The smaller the buffer, the more precise is the landscape definition that we know for sure the yellow fever virus (YFV) can circulate in. On the other hand, we may (1) incorporate noise due to location error (from GPS or reporting errors); and (2) leave out other environmental characteristics that are also probably favorable to YFV circulation, given that infected mosquitoes and NHP move around. Although there exist studies regarding the estimation of YF-related biotic parameters such as host movement and flight radius of vectors [13, 14], they tend to be excessively coarse (e.g. 11 km for vectors [8]) for the purpose of fine-grained modeling. Thus, to mitigate the injection of arbitrary environmental characteristics into the modeling (noises), we decided to limit the analysis to the maximum buffer of 1000 m. In this sense, the 100 m buffer provides the finest analysis scale, 500 m the middle term, and 1000 m the coarsest scale. Since there is no proven optimal buffer size for this kind of study, the multi-buffer strategy should lessen the impact of that uncertainty while making the models more reliable.

In general, there will be a spatial characterization for each time point, resulting in multiple characterizations for a given region. The total number of spatio-temporal characterizations will depend on the temporal resolution of the layer: some are considered static/atemporal (such as altitude) whereas others, such as rainfall and temperature, have monthly resolution.

To produce a prediction map (heat map) from the trained ensemble model so that suitable areas for yellow fever can be revealed, we first generate a regular grid of points over the map, and then characterize them against the most recent environmental layers. This is the typical case, and leads to a near-present snapshot of YF suitability—although not covered in this research, future projections of disease suitability require themselves future projections of the input environmental data. To make it feasible to compute predictions for the entire country of Brazil, which has continental dimensions, we set the grid resolution to 1 km, resulting in 7,110,036 points—working at 30 m would lead to ∼8 billion points, and would be practical only for subnational levels, such as individual states and municipalities.

### 2.5 Merging outputs: concatenating and joining Concatenate

The characterization procedure from the previous section results in a bunch of intermediate files, each corresponding to an input environmental layer at a given time period. Furthermore, due to the parallel execution needed to tackle huge amounts of points—where each computing core processes independently a partition of the points set—additional output files are generated. All scattered files related to a given environmental layer are concatenated together, and a field named *datetime* is included to register the time period of the characterization.

*Join*. Once we have the spatio-temporal characterization for all environmental layers, we need to link each point—positive case or grid point—to the nearest buffer characterization time-wise. This ensures that the retrieved environmental context is the best possible description of the context when the virus circulated in the region. This is a prerequisite for capturing the seasonality of the disease. Also, the joining procedure merges the variables from all environmental layers, effectively creating a complete (raw) dataset for each buffer size. Each per-buffer dataset ended up with 918 statistical environmental features (plus a geocode field), which is a product of 51 classes by 18 descriptive statistics. The next section describes how these features are filtered out so that machine-learning methods can be properly applied.

### 2.6. Filtering out features / feature selection

By their very nature, the calculated descriptive statistics tend to be highly correlated among them. For instance, the *normalized* statistics, although they enable inter buffer comparisons, are merely the original statistics divided by the buffer area (a constant). Machine-learning methods are typically sensitive to feature correlation, thus removing correlations is crucial for their performance [15]. On top of that, it is well-known in the literature [16] that the ratio between the number of training instances (*n* = 545) and the number of features (*n* = 918) should be at least greater than one so the model has more data points than variables to estimate, which currently is not the case (545/918 = 0.59).

### Low-variability features

The first step in feature selection is to filter out the constant or near-constant features, which therefore do not contribute significantly information-wise. This is achieved through the calculation of *coefficient of variation, CV* = *σ*/|*µ*|, where *σ* and *µ* are the standard deviation and mean, respectively [17]. All features with *CV* < 0.05 were removed, as such features are unlikely to provide meaningful information for the model. For buffer sizes 100 m, 500 m, and 1000 m, this procedure alone eliminated respectively 265/918 (29%), 245/918 (27%), and 211/918 (23%) features.

The computation of the coefficient of variation was solely performed on the training datasets, individually for each buffer size. One can notice that as the buffer size increases, less likely it is for a certain feature to have low variability; this is due to the larger influence area which tends to aggregate more information (or noise).

### Multi collinear features (VIF)

The last step regarding feature selection was to apply the Variance Inflation Factor (VIF) method [18], which filters out features with high multicollinearity. This is an iterative procedure in which at a given iteration every feature has its multicollinearity measured by VIF = 1/(1− *R*^2^), with *R*^2^ being the coefficient of determination; the most multicollinear feature is removed, and the iterations continue until the degree of multicollinearity (VIF) of every remaining feature is no more than a threshold. The frequently suggested threshold value of 5 was used [19].

The VIF method was applied to each buffer size over the training sets, after filtering out the low-variability features as described before. As a result, only 49, 65, and 72 features were kept for buffer 100 m, 500 m, and 1000 m respectively. Since there was divergence among buffer sizes, we decided to union the selected features of all three buffers, which finally led to 140 features. Table 1 breaks down the selected features aggregated by environmental layer. Since features were selected from every layer, this means that they are singular information-wise and thus they can all potentially contribute to model YFV occurrence.

**Table 1:** Selected features aggregated by layer.

| Layer/class | \|selected\| | percentage |
| --- | --- | --- |
| Altitude (SRTM) | 5 | 27.78% (of 18) |
| Rainfall (CHIRPS) | 5 | 27.78% (of 18) |
| Temperature max (CHIRTS) | 3 | 16.67% (of 18) |
| Temperature min (CHIRTS) | 2 | 11.11% (of 18) |
| Climate normals (WorldClim) | 30 | 8.77% (of 342) |
| Land use & cover (MapBiomass) | 95 | 18.85% (of 504) |

### 2.7 Training & predictions

At this stage we have 545 points and their accompanying environmental features. Strictly speaking, the goal is to model the underlying distribution of these values so that we can measure the similarity of any arbitrary point (its environmental characterization) w.r.t. this distribution, to ultimately predict whether they are *inlier* (suitable for yellow fever) or *outlier* (not suitable).

Several data-driven approaches can address this task, ranging from simple envelope methods—which estimate the underlying data distribution without explicitly modeling it—to statistical techniques and, ultimately, more sophisticated machine learning methods. Due to its proven capability of tackling complex problems in a wide variety of domains as well as its long-standing popularity, we employed here the Support Vector Machine (SVM) method, more precisely its one-class variation (One-Class SVM) [20] since we are dealing with presence-only data—in Brazil, at least, data about truly unsuitable areas for yellow fever is very scarce and therefore would not be useful for modeling.

The One-Class SVM is an unsupervised algorithm used for anomaly detection—in our case, *anomaly* can be understood as *not suitable* for yellow fever. It learns a decision boundary that encloses the majority of the data, identifying outliers as points falling outside this boundary. The model uses a kernel function, typically the radial basis function (RBF), to capture non-linear patterns. Two key parameters control its behavior [21]:

- *v* (0 < *v*≤ 1)This parameter serves as an upper bound on the fraction of training errors (i.e., the proportion of positive points allowed to lie outside the decision boundary) and a lower bound on the fraction of support vectors. It directly controls the trade-off between model sensitivity and robustness to outliers. A smaller value of *v* leads to a tighter decision boundary (fewer training false negatives), while a larger *v* increases model tolerance for contamination (noise) in the dataset.
- γ (γ > 0) This is the kernel coefficient for the RBF kernel, controlling the influence of individual training examples. A small γ implies a larger similarity radius (more influence from each point), resulting in a smoother decision boundary. In contrast, a larger γ makes the model more sensitive to small-scale patterns, which can lead to overfitting.

This method is effective when training data contains mostly normal (“positives”) instances and labeled anomalies (“negatives”) are rare or absent, which fits well our problem since our training dataset is composed of positives-only data.

Determining the optimal parameter set in presence-only problems (generative models) is typically difficult. To work around this limitation while further improving prediction robustness, we employ an ensemble model, comprised of sub-models built from different sets of parameters. In addition to *buffer size* and SVM’s *v* and γ, we also took into consideration the parameters of Principal Component Analysis (PCA [22]), which we adopt as a pre-processing step, in particular the amount of *explained variance*.

The ensemble is formed by the Cartesian product of selected range of arguments from buffer size, *v*, γ, and PCA; respectively: 100 m, 500 m, 1000 m ×{90%, 50%, 10%, 5%, 1%, 0.5%, 0.01%, 0.005%, 0.001%}×{scale, auto, 0.9, 0.5, 0.1, 5e −2, 1e −2, 5e −3, 1e −3, 5e −4, 1e −4}×{none, 90%, 95%, 99%} = 3× 9× 11× 4 = 1188 members. We deliberately chose a wide range of arguments per parameter, including their usual values, to ensure an extensive coverage of the parameter space.

A side effect of this relaxation is that some combinations result in submodels that exhibit poor training accuracy, meaning that they were not able to effectively learn how to separate *inliers* from *outliers*. To prevent underperforming submodels from degrading the overall ensemble performance, we discarded those whose training accuracy were below 80%, a parameter that we called *training accuracy threshold* (τ). In practice, we are assuming that the training datasets have at maximum 20% of impurity (such as false positives and data noise). This post-processing shrunk the ensemble model roughly by half, from 1188 to 532 members. Moreover, we also repeated all experiments with τ = 90% (ended up with 359 members) to analyze its impact on ensemble model performance (see Section 4.1).

### Ensemble statistics

For each grid point we make *n* predictions, with *n* equal to the number of ensemble members. This is a time-consuming process which requires almost 3.8 billion model evaluations (7110036×532) — to speed up this phase, we ran it in parallel on an Intel Xeon machine with 48 cores at 2.4GHz and 768GB RAM. The 532 predictions per grid point were aggregated into statistics according to the following three categories: *overall, positive*, and *negative*. The first considers the entire range of model predictions for calculating descriptive statistics, which can be either positive (above zero value) or negative (below zero). The *positive* category only takes into account predictions above zero; analogously, *negative* only takes into account predictions below zero. For each category, the descriptive statistics shown in Table 2 are calculated.

### 2.8 Presentation of the predictions

From the ensemble, we present the results to decision makers as suitability maps for Yellow Fever. There are many ways the information can be conveyed:

1. **Overall confidence** [0, 1]: how certain is the (ensemble) model at each region (grid point). This effectively measures the agreement level of the submodels, either towards positiveness or negativeness, and is calculated as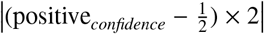 Based on that, one could use this metric to filter out predictions below a given confidence threshold due to their uncertainty.
2. **Positive confidence** [0, 1]: how certain the model is about the region being positive. It is the proportion of submodels that predicted a positive suitability; thus, the higher this value, the more suitable is the region for disease occurrence. This metric is the one we deem most adequate in general.
3. **Positive count** ×**positive mean** [0, ∞]: intensity of positiveness. While *positive confidence* discards the individual degree of suitability of the submodels, multiplying the number of positive predictions by the corresponding mean suitability provides the total intensity of suitability. The downsides are the difficulty to compare different suitability maps (due to the open interval), and sensitivity to anomalies.
4. **Aggregated municipality-based maps** [0, 1]: In decision making, aggregated suitability maps sometimes are more informative than fine-grained ones. For this, we provide two municipality-based aggregation levels: (1) averaged *positive confidence*; and (2) simplified three-stage average with *low, medium*, and *high* suitability/priority—the intervals are respectivelly (0, *P*_33.3_], (*P*_33.3_, *P*_66.7_], and (*P*_66.7_, 1], with *P*_*k*_ denoting the *k*-th percentile.
5. **Varying sensitivity** [0, ∞]: finally, thanks to the ensemble modeling, it is straightforward to produce suitability maps of varying sensitivity, which can help to prioritize areas for disease prevention/mitigation. This is accomplished by retrieving the following statistics: *min* (lowest), *P*_10_ (very low), *P*_25_ (low), *median* (moderate), *P*_75_ (high), *P*_90_ (very high), *max* (highest).

## 3. Results

The map depicted in Figure 4 represents the predicted environmental suitability for yellow fever for each grid point, according to the degree of positiveness of the ensemble model (*positive confidence*). The range of values lies in [0, 1] since the metric is the percentage of ensemble members (submodels) that indicated the region as suitable.

**Figure 4:**
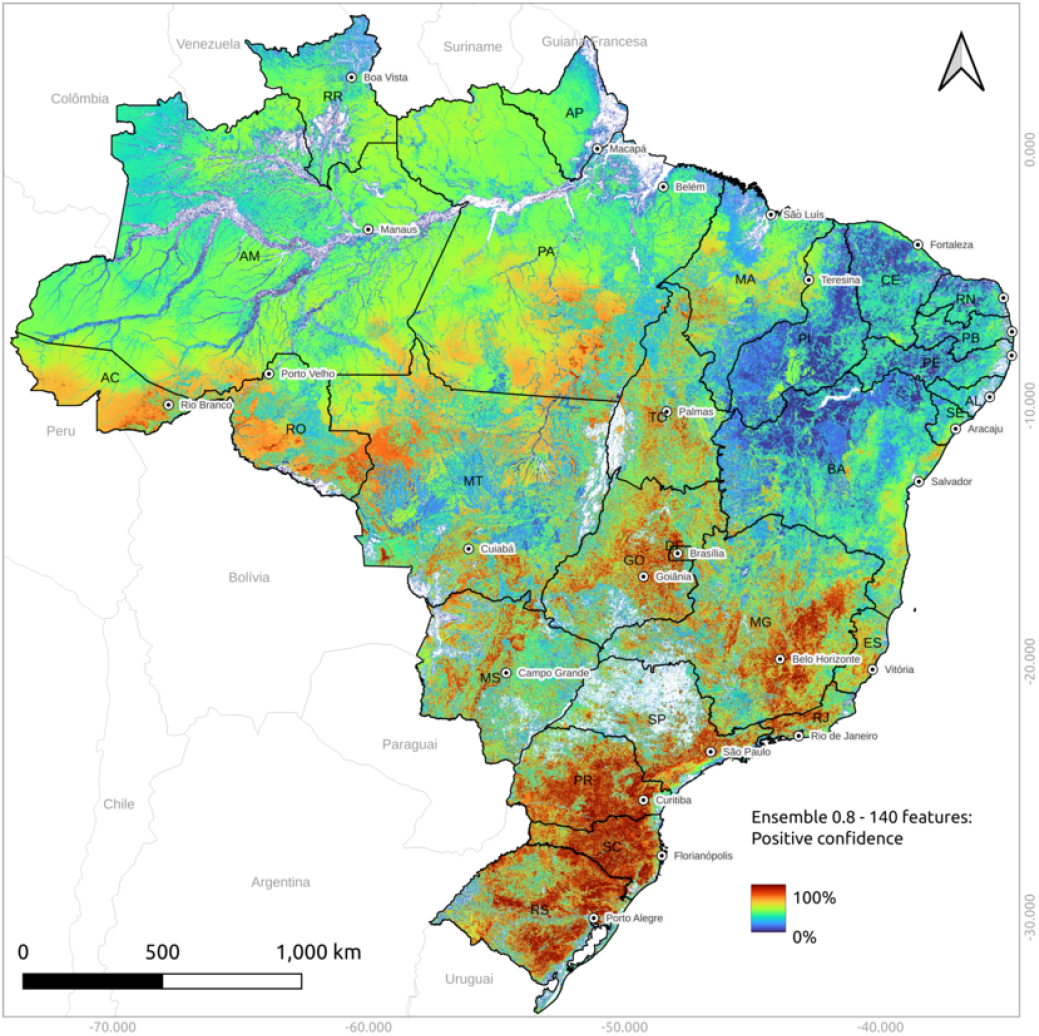
Fine-grained prediction map of yellow fever environmental suitability as the degree of positiveness of the ensemble model.

Southern Brazil presents the highest mean suitability level (MSL), 0.64, strongly influenced by the distribution of cases in the region used as training instances (434/545 = 79.6%). The region contains, across its three states, the largest and most extensive clusters of strong suitability for YF in the country. In the state of Rio Grande do Sul (RS), high-suitability areas are concentrated in the northeast, along the border with Santa Catarina). They also extend southeast and reappear in the center-east (Porto Alegre region), with smaller strips in the center-west and northwest (northern portion of the border with Argentina along the Uruguay River). In Santa Catarina state (SC), elevated suitability is observed in the center-south, particularly in the south-eastern portion bordering RS. Additional suitable zones occur in the north (border with Paraná), and around the center-east (Florianópolis region), with smaller patches appearing in the center-west. In Paraná (PR), the main zones of pronounced suitability are located in the center-south and the east-southeast (Curitiba region). Smaller corridors of suitability are found in the west, southwest, and northeast. The Southeast region comes next, with MSL of 0.46, even though it accounted for only 9.5% of the training cases (52/545). The region displays extensive regionalized areas of elevated suitability across its four states. In the São Paulo state (SP), the most suitable areas are in the southeast (Capital region), the south (near the southern coast), the SP–RJ axis (Vale do Paraíba), and smaller clusters in the center-south and northeast. In Minas Gerais (MG), suitability is notable in the center (Belo Horizonte region) and in the east (Zona da Mata–border with RJ–and parts of the northeast of the state, connecting with ES and southern BA), with additional localized clusters in the north, northwest, and southwest. In the state of Rio de Janeiro (RJ), the highest suitability occurs along the border with MG (Zona da Mata) and SP, with additional strips in the Capital region and in the eastern coast. In Espírito Santo (ES), suitability is concentrated in the central and northwestern portions of the state. In close third, with MSL of 0.44, came the Central-West region. The region contains a mosaic of high and medium suitability zones and corridors across its three states. In Goiás (GO), clusters are concentrated in the center (Goiânia region) and the east (adjacent to DF), with additional clusters in the north and northwest. In Mato Grosso do Sul (MS), suitability is high in the center-north (Campo Grande region) and intermediate in restricted strips in the west (southern Pantanal). In Mato Grosso (MT), intermediate suitability predominates in the south (Cuiabá region and border with MS). A small cluster of strong suitability occurs in the southwest (near the border with Bolivia) and intermediate portions along the border with RO. The North region, with MSL of 0.39, ranks fourth. Although the region is well known as endemic for yellow fever [23], only seven cases were reported during the study period. One reason for the surprising under-reporting has to do with the historically high vaccination coverage in the region [24], which tends to reduce the intensity of surveillance. Another reason is the difficulty in accessing regions to collect samples of dead NHP for laboratory analysis. Despite this data scarcity—only 1.3% of the training data—the ensemble model was able to recognize intermediate suitability areas in four states: in Rondônia (RO), in the southeast and parts of the center-west extending close to the Bolivian border; in Acre (AC), in the center-west reaching areas near the Peruvian border; in Pará (PA), in the center-south; and in Tocantins (TO), with central clusters of notable suitability. The Northeast region, which had only one reported case during the study period, showed the lowest MSL (0.28), which comes as no surprise due to the lack of data. Still, small zones of moderate suitability were identified in two states. In Maranhão (MA), they are concentrated in the central and western portions; and in Bahia (BA), in the south and coastal areas (where fragments of Atlantic Forest persist). In the other states—Piauí (PI), Ceará (CE), Rio Grande do Norte (RN), Paraíba (PB), Pernambuco (PE), Alagoas (AL), and Sergipe (SE)—low suitability predominates, with very localized and restricted intermediate clusters. The suitability map we have just discussed (Figure 4) should be interpreted with the aid of the prediction confidence map in Figure 5, which in practice represents the agreement level of the ensemble model. The more yellow the region, the more reliable the prediction—either as suitable or unsuitable; conversely, blue areas indicate high uncertainty. In the North region of Brazil, for instance, the model is very confident about the suitability level of the immense Amazon and Negro rivers, which according to Figure 4 are, unsurprisingly, highly unsuitable for yellow fever. Except for these rivers and other few spots, the model exhibits low confidence about the predictions in the region, meaning a lack of knowledge about its environmental characteristics, mainly a consequence of data scarcity. Another instance of pronounced confidence is the northern portion of the state of São Paulo (SP), which is largely dominated by sugarcane plantations and, as such, does not generally sustain high environmental suitability for yellow fever. However, small forest remnants in municipalities such as São José do Rio Preto and Ribeirão Preto, where cases were reported, are captured by the model as localized clusters of higher suitability embedded in this otherwise low-risk sugarcane matrix. On top of assisting in the interpretation of predictions, the confidence map can also be used as a mask to filter unreliable predictions out (e.g. hiding all predictions below 90% confidence), thus allowing decision makers to concentrate on the most trustworthy ones.

**Figure 5:**
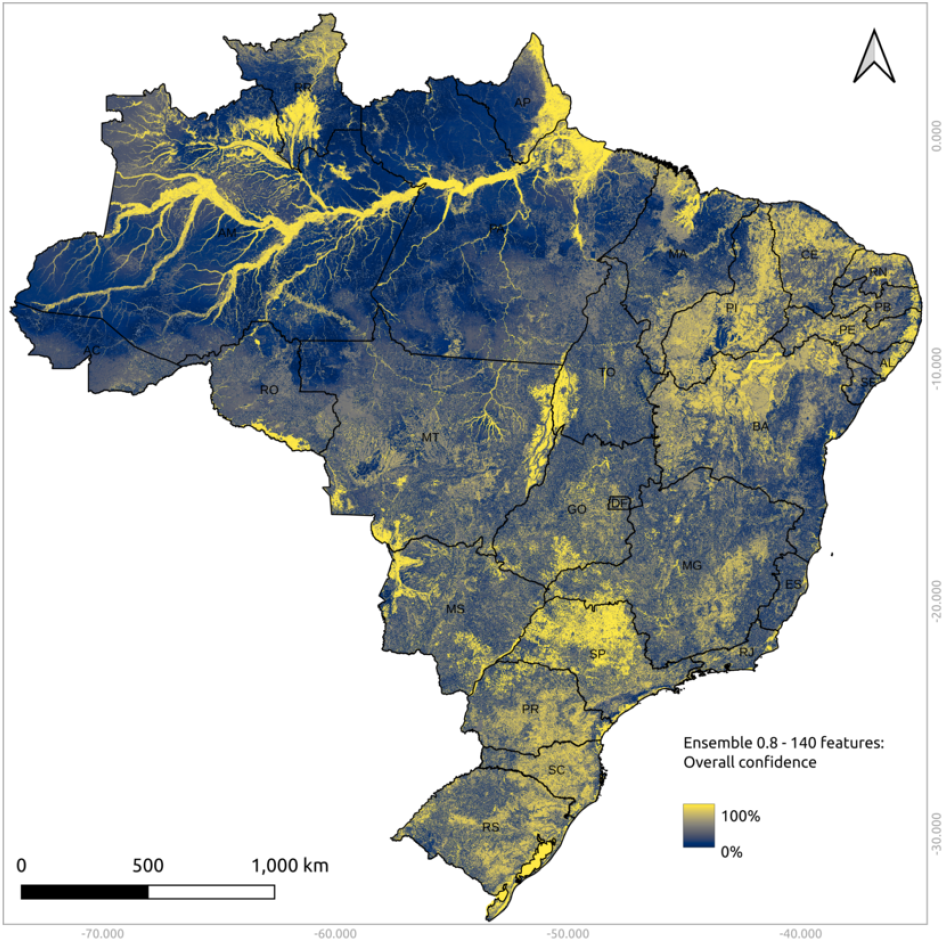
Overall prediction confidence map (ensemble agreement).

As mentioned in Section 2.8, to facilitate communication with decision makers and to serve the country’s political administrative divisions, predictions may also be aggregated. Figure 6 shows the suitability map averaged per municipality (mean value), whereas the simplified three-stage map is presented in Figure 7. Although they lose the fine-scale of predictions, they are typically helpful for conveying the overall picture for the purpose of decision making, such as vaccination prioritization [25].

**Figure 6:**
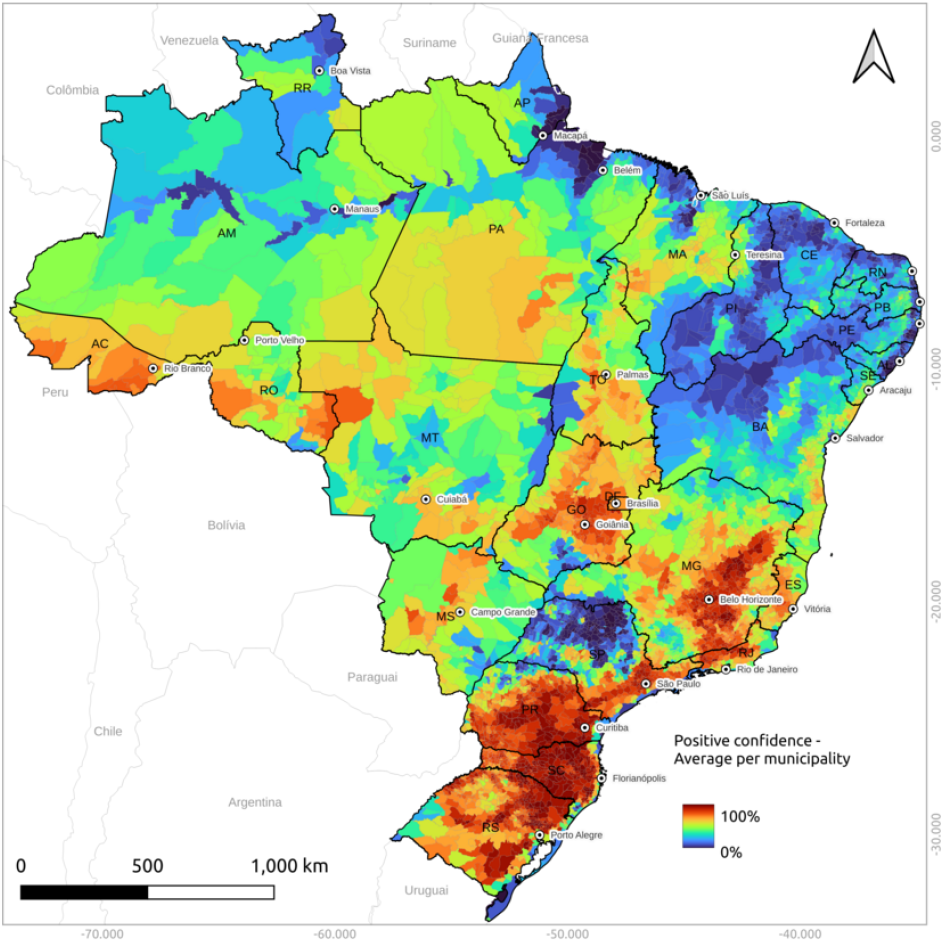
Averaged per-municipality prediction map of yellow fever environmental suitability as the degree of positiveness of the ensemble model.

**Figure 7:**
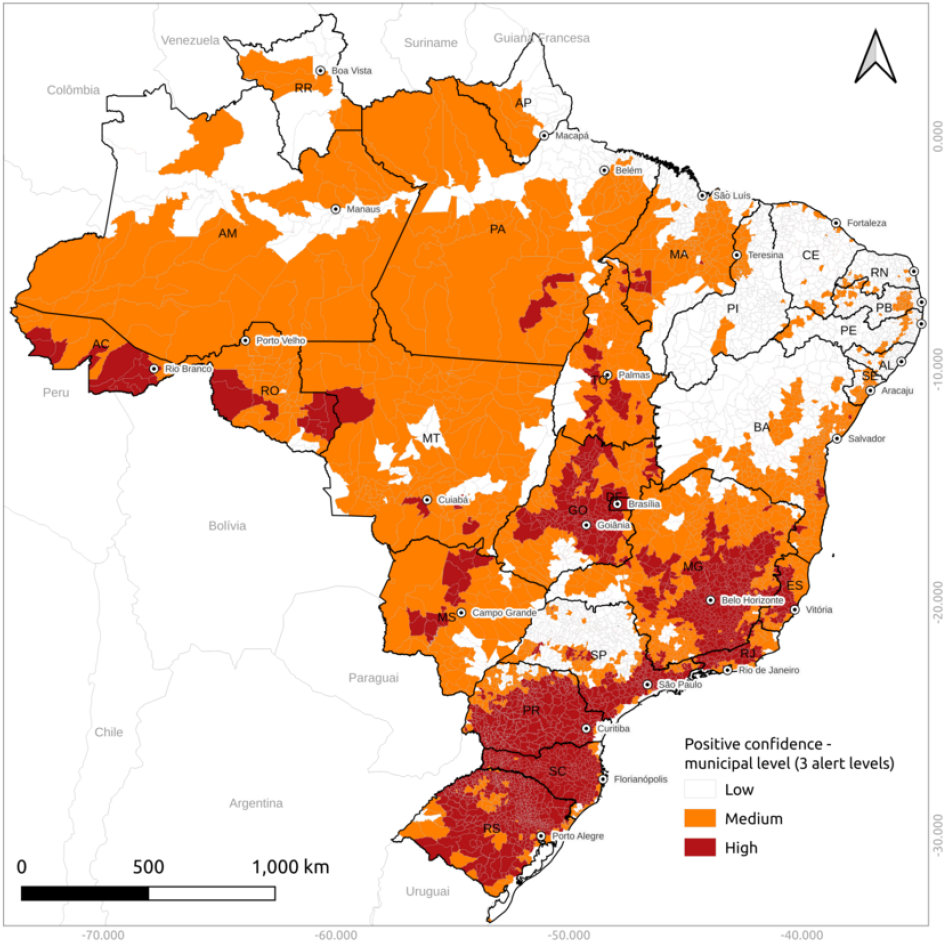
Three-stage averaged per-municipality prediction map of yellow fever environmental suitability as the degree of positiveness of the ensemble model.

Thanks to the ensemble model, it is straightforward to control the trade-off between sensitivity and precision. For a given region, taking the maximum predicted suitability level across all submodels produces the most sensitive ensemble model; conversely, the minimum level leads to the most precise model. While the former tends to be over-alarmist, potentially generating numerous false positives, the latter may be excessively conservative, misclassifying truly positive areas as unsuitable for yellow fever. Relying on extreme values, however, impacts negatively the robustness of the ensemble model, being therefore rarely adopted in practice. Instead, it is preferable to use more reliable alternatives such as percentiles. Figure 8 illustrates four maps of varying sensitivity, obtained by post-processing the ensemble model: 10%, 25%, 50%, and 75% percentile. The more the sensitivity, the larger the suitability area; in other words, a map of a higher sensitivity expands the area of a map of lower sensitivity. Which sensitivity threshold to use for surveillance is up to the decision maker and depends on many factors such as resource availability, scale of analysis, and false-positive versus false-negative rate acceptance. As a general rule, more precise predictions (less sensitivity) are preferable when false positives are an issue, whereas more sensitiveness are preferable in contexts where false negatives have serious consequences.

**Figure 8:**
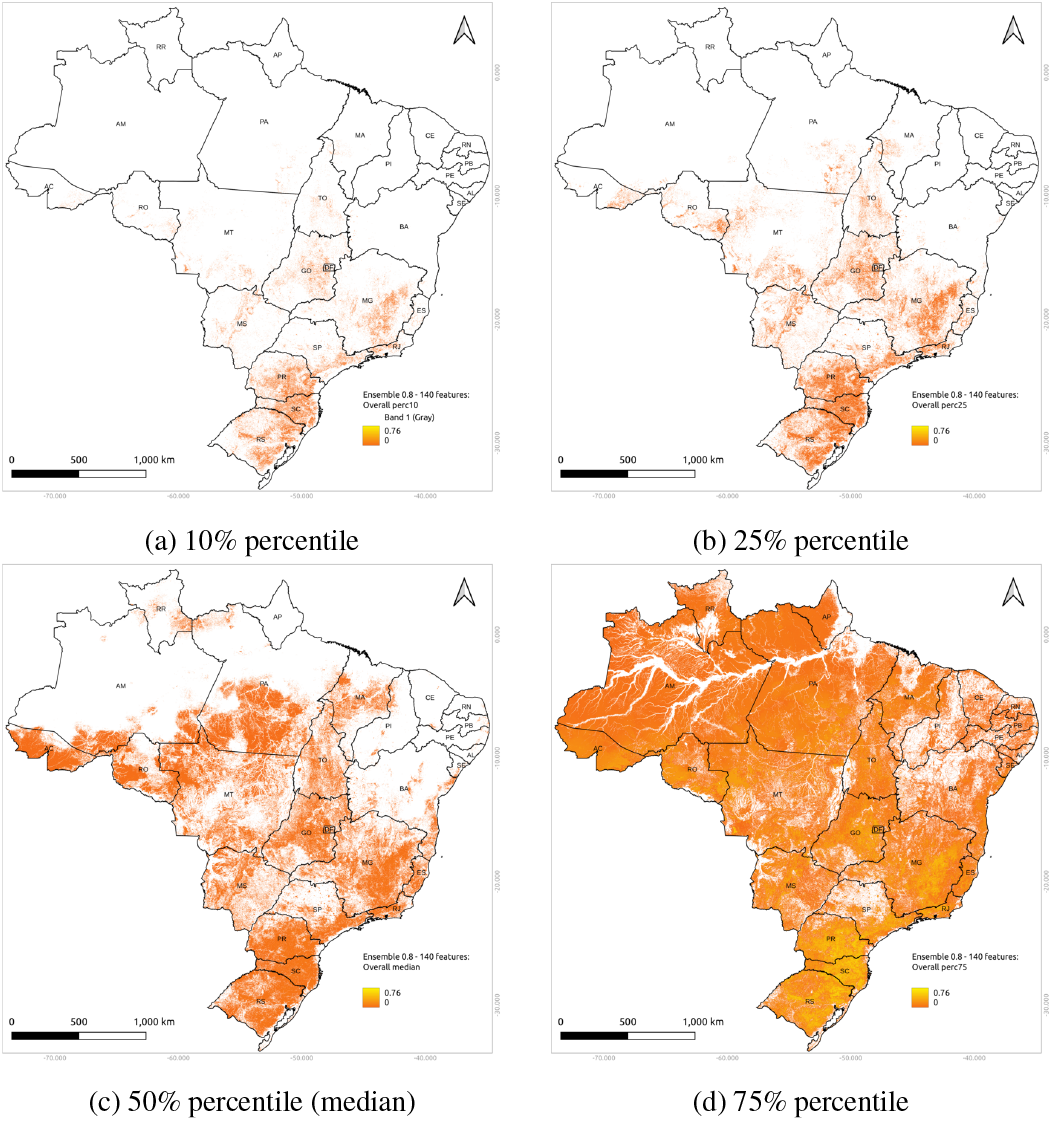
Maps of varying sensitivity: 10%, 25%, 50%, and 75% percentiles. The lower the percentile value, the higher the priority of the area is for surveillance (higher “risk”).

### 3.1. Validation

We already know that our ensemble model performs with at least 80% accuracy on the training cases dataset, due to the fact that this threshold was a pre-requisite of the ensemble model formation (refer to Section 4.1 for a comparison with a tighter threshold). A more meaningful feature, though, is the model’s ability to perform better on unseen regions, that is, to anticipate yellow fever occurrence. To assess the generalization ability, we (i) trained the ensemble model with YF cases up to the year of 2024 (see Figure 1), (ii) performed the prediction for the whole country (Figure 4), and finally (iii) measured the suitability level of each region that intersects each of the 23 new YF cases in 2025. As shown in Figure 9, all validation cases occurred in regions highly suitable for yellow fever. The highest predicted suitability was 97%, whereas the lowest was 58%, which is still expressive and means that the ensemble model correctly classified all validation cases. The overall suitability was 74%.

**Figure 9:**
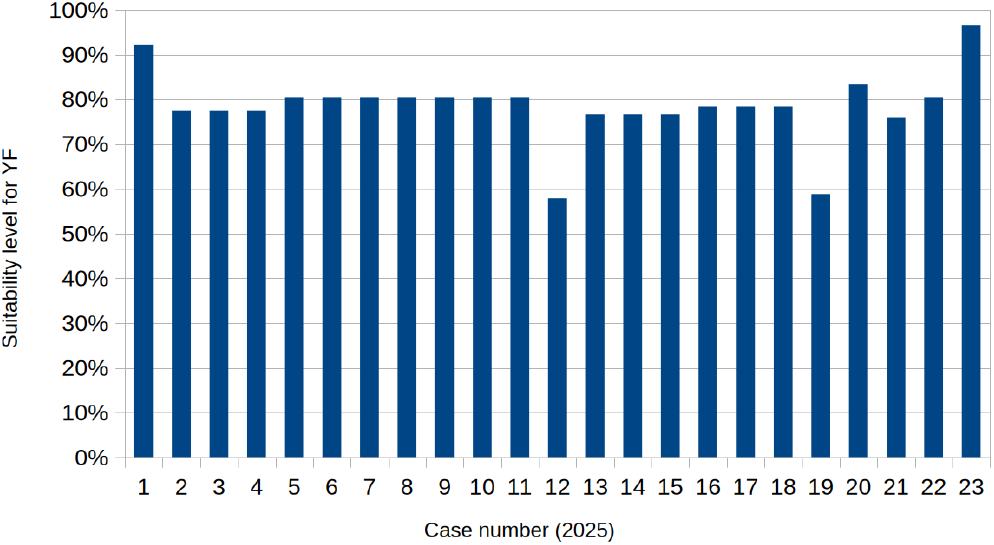
Suitability level for each validation case (year of 2025).

### 3.2 Feature analysis

Understanding the complex interplay of factors influencing yellow fever dynamics is crucial for effective public health interventions, disease forecasting and prevention. Moreover, it supports the validation of the model by specialists as well as its acceptance by decision makers. While an ensemble of SVM models is known to perform well on challenging problems, its many parameters and nonlinear interactions make it difficult to determine the precise contribution of individual input features to the model’s predictions.

This section introduces the application of SHAP (*SHapley Additive exPlanations*) [26] analysis to elucidate the drivers behind the output of our yellow fever geospatial model. SHAP is a game-theoretic approach that analyzes how changes in input features affect model predictions, allowing for insights into feature importance and model behavior. The application was carried out directly on the trained ensemble model, which was built from a subset of the original features (see Section 2.6).

The first analysis is presented in Table 3, where the overall feature contribution aggregated by layer is quantified in terms of SHAP values. We observe that *Land use and over* accounts for the largest influence in the model regardless of the metric. *Climate normals* ranks second w.r.t. the sum of SHAP values, but this may be biased due to the fact that it involves the second major group of selected features (Table 1). Instead, when the mean SHAP value is analyzed, *Temperature* takes the second place, followed by *Altitude* and then *Rainfall*, and finally *Climate normals* as the least important.

**Table 2:** Ensemble statistics.

| statistic | meaning |
| --- | --- |
| *mean* | mean prediction value (moderate sensitivity) |
| *min* | minimum prediction value (lowest sensitivity) |
| $P_{10}$ | percentile 10 (very low sensitivity) |
| $P_{25}$ | percentile 25 (low sensitivity) |
| $P_{50}$ | median (moderate sensitivity) |
| $P_{75}$ | percentile 75 (high sensitivity) |
| $P_{90}$ | percentile 90 (very high sensitivity) |
| *max* | maximum prediction value (highest sensitivity) |
| *std* | standard deviation |
| *confidence* | agreement level of the ensemble members [0,1] |
| *count* | number of ensemble members |

**Table 3:** Overall impact of features aggregated by layer.

| layer | \|SHAP value\| ($\times 10^{-2}$) | | |
| --- | --- | --- | --- |
|  | sum | mean | max |
| Land use and cover | 35.6 | 0.375 | 0.722 |
| Climate normals | 9.48 | 0.316 | 0.367 |
| Temperature | 1.84 | 0.367 | 0.411 |
| Altitude | 1.82 | 0.364 | 0.376 |
| Rainfall | 1.73 | 0.346 | 0.352 |

Regarding the individual contribution of the classes belonging to *Land use and cover*, the ten most influential ones from the model’s perspective are plotted in Figure 10. Section 4.2 presents an in-depth discussion of these most impacting classes in the context of environmental suitability as predicted by the model.

**Figure 10:**
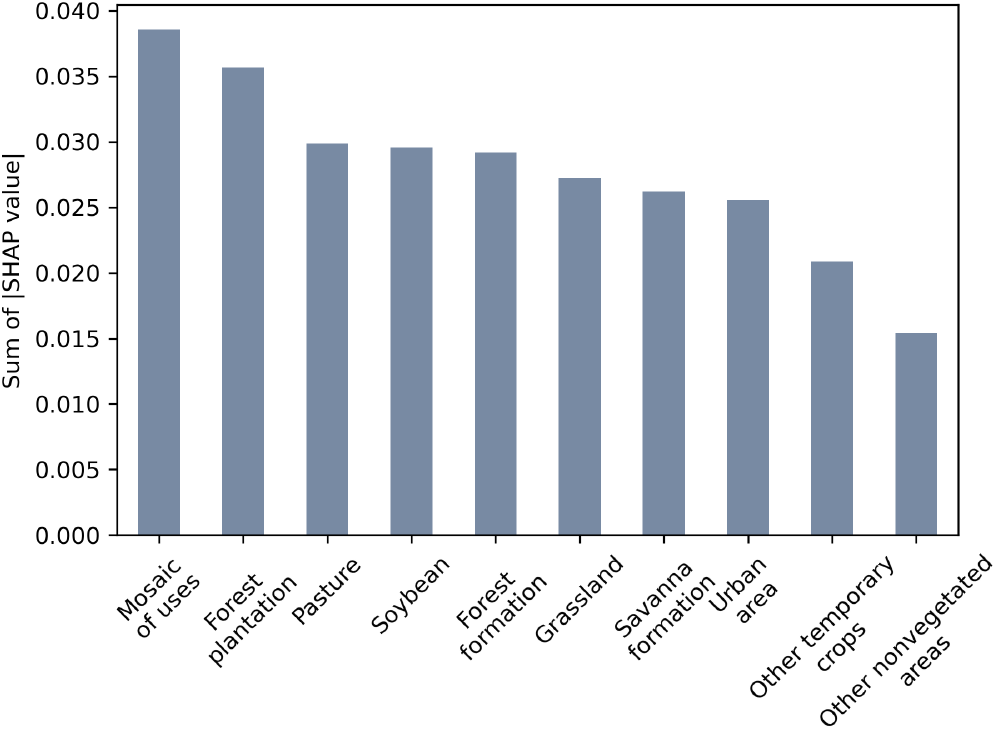
Overall impact of *Land use and cover* classes.

## 4. Discussion

In the following sections we discuss two main aspects of the methodology and results. First, we examine the implications of adopting a different threshold for training accuracy, which directly affect model calibration and predictive performance. Second, we analyze the environmental characteristics identified by the model as relevant for yellow fever occurrence (by means of SHAP analysis), providing insights into ecological patterns that may guide surveillance and prevention strategies.

### 4.1 Training accuracy threshold (τ)

For the training of ensemble model described in Section 2.7 we used a parameter that controls the required minimum accuracy on the training dataset for a submodel to be part of the final ensemble, the parameter τ. Although we opted to use τ = 80%,the question remained whether a stricter value would be better. Figure 11 shows that the threshold τ = 90% yields very similar spatial patterns to τ = 80%, just with an increased intensity (29% higher on average). In fact, the Pearson correlation is ρ = 0.99, meaning that the behavior is effectively the same.

**Figure 11:**
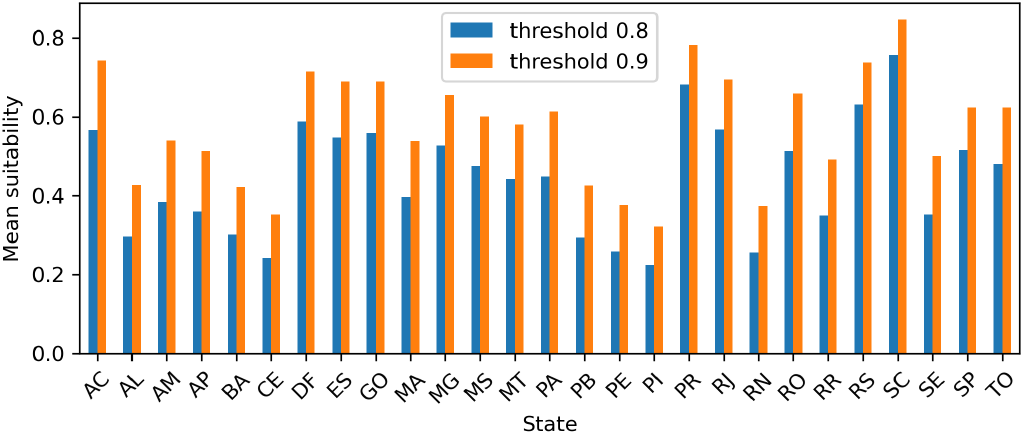
Ensemble model’s predictions comparison with τ = 80% and 90%.

### 4.2 Analysis of environmental characteristics

For this analysis we split up the country-level predictions (Figure 4) into two antagonistic groups: low and high mean suitability level predictions, *L*_MSL_ and *H*_MSL_ respectively, aiming at analyzing the environmental discrepancies between them. The *L*_MSL_ group consists of all predictions that fall into the (0, *P*_33.3_] interval, with *P*_*k*_ denoting the *k*-th percentile. Conversely, predictions within the percentile interval (*P*_66.7_, 1] are assigned to the *H*_MSL_ group. We purposely threw out predictions lying in (*P*_33.3_, *P*_66.7_] to lessen the effect of uncertainty on the analysis. Then, based on SHAP’s feature ordering by importance as described in Section 3.2, we compare the percentage of change of each feature’ mean value in *H*_MSL_ relative to its mean value in *L*_MSL_, effectively measuring the extent of variation which might be associated with the yellow fever occurrence.

On average, *H*_MSL_ regions contain 21% less *mosaic of uses*, 82% less *forest plantation*, and 36% more *pasture*. They have almost no *soybean* (−99%), but 184% more *forest formation* and 8% more *grassland*. In contrast, they include 92% less *savanna formation*, 346% more *urban area*, 8% less *other temporary crops*, and 92% less *other non-vegetated areas*.

The largest positive differences were found in *urban area* (+346%) and *forest formation* (+184%). Although it is difficult to assess the sampling bias influence in these findings, especially regarding the urban area difference where proximity to urban infrastructure increases detection and reporting of NHP carcasses, they may also indicate that the disease is focal in fragmented, peri-urban and forest patches rather than in remote continuous forest. In contrast, *L*_MSL_ regions contain much more *soybean* area (+14335%), *other non-vegetated areas* (+1236%), *savanna* (+1137%), and *planted forests* (+468%). They also feature 27% more *mosaic of uses* and just 9% more *other temporary crops*. These correspond to open, homogeneous landscapes with little humid forest or wetlands, less favorable to the sylvatic cycle.

Overall, higher suitability is associated with humid forests, riparian forest corridors, and agroforestry mosaics, while lower suitability is linked to extensive open agriculture with reduced habitat diversity and connectivity.

## Conclusions

Sylvatic yellow fever is a transmissible disease of high lethality that exhibits intricate dynamics, involving human and non-human primates, mosquitoes, environment, and the virus itself. The disease can even devastate entire populations of nonhuman primates and kill humans, even though a vaccine for it has existed for decades. On the other hand, epidemiological, ecological, and spatial models can provide valuable tools for anticipating outbreaks, guiding targeted vaccination strategies, optimizing vector control, and informing early warning systems.

In this study we presented a framework for large-scale geospatial modeling of YF occurrence in Brazil, which historically has been the country most affected by the disease worldwide. To the best of our knowledge, this is the first fine grained global modeling of YF in Brazil. On top of having a methodology capable of tackling such large scale modeling, it also requires a huge amount of computational resources as well as specialized high-performance software tools to leverage them. Built on precisely located disease cases, we employed a comprehensive set of spatiotemporal high-resolution environmental characterization at multiple scales together with an ensemble of machine-learning models for robust predictions. We demonstrated different ways of analyzing the generated prediction maps and estimated the impact of individual feature classes on model output through SHAP analysis, showcasing how we can work on model interpretability. Then, we followed with a quantification of the environmental differences between high and low suitable regions for YF as a preliminary effort to reveal possible key drivers of YF occurrence.

While this study represents a step forward in the field of YF modeling, several avenues for future research remain. First, the effect of sample bias could be mitigated by adopting a cluster based case weighting scheme in which nearby cases would share their influences on model training. Second, the incorporation of fragmentation metrics as additional environmental variables would hopefully improve model accuracy as there exists evidence linking forest fragmentation to YF occurrence [27, 28, 29]. Third, investigations into the spatial extent of landscape effects on YFV circulation—according to host/vector ecology, spatial resolution, model sensitivity, and location errors—may contribute to the determination of optimal buffer sizes. Fourth, exploring the integration of vaccination coverage into the modeling would be helpful in risk assessment for humans. Finally, developing specific models per NHP genus could refine the overall accuracy given that their ecological parameters differ between genera.

## Data Availability

The data used in this study were obtained from the Brazilian Ministry of Health and contain georeferenced information on reported cases of sylvatic yellow fever.
Due to the presence of sensitive geographic coordinates that could potentially allow identification of case locations,
these data are not publicly available and cannot be shared by the authors.
Access to the data is restricted and may be granted only upon formal request and authorization by the Brazilian Ministry of Health.

## Data Availability

The data used in this study were obtained from the Brazilian Ministry of Health and contain georeferenced information on reported cases of sylvatic yellow fever. Due to the presence of sensitive geographic coordinates that could potentially allow identification of case locations, these data are not publicly available and cannot be shared by the authors. Access to the data is restricted and may be granted only upon formal request and authorization by the Brazilian Ministry of Health.

The computational workflow and code used for the analyses described in this study are available from the corresponding author upon reasonable request.

## APPENDIX A Environmental layers’ maps

**Figure A.12:**
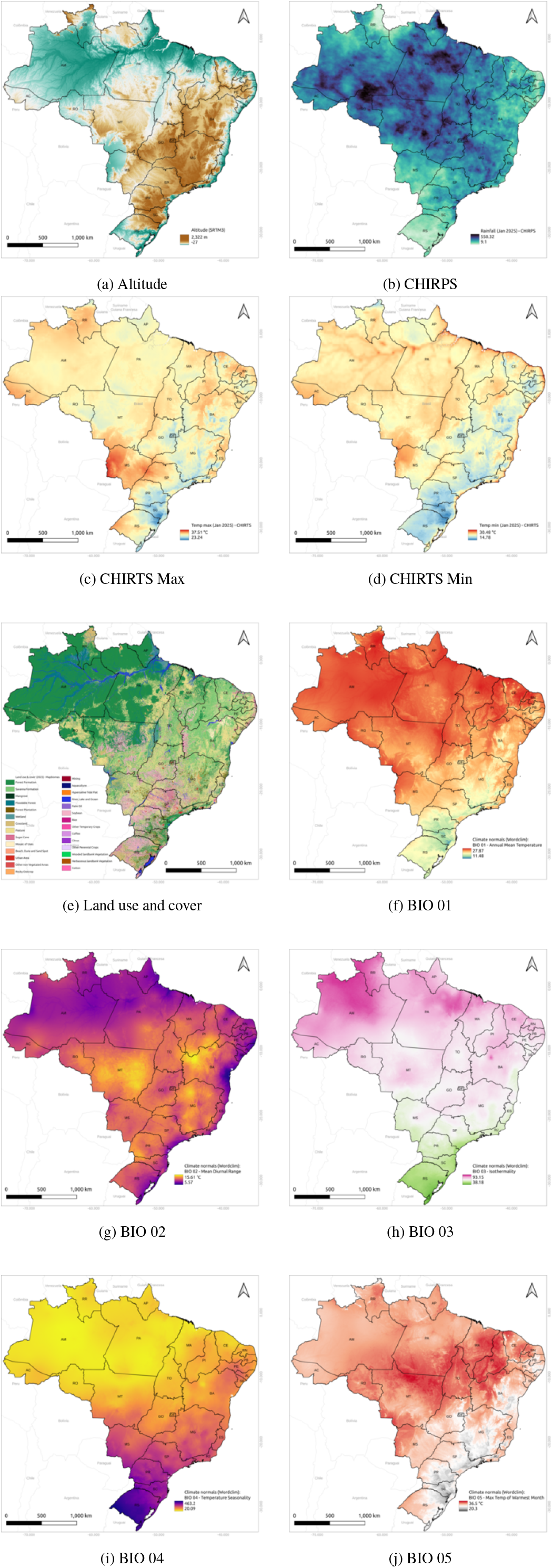
Environmental layers (I)

**Figure A.13:**
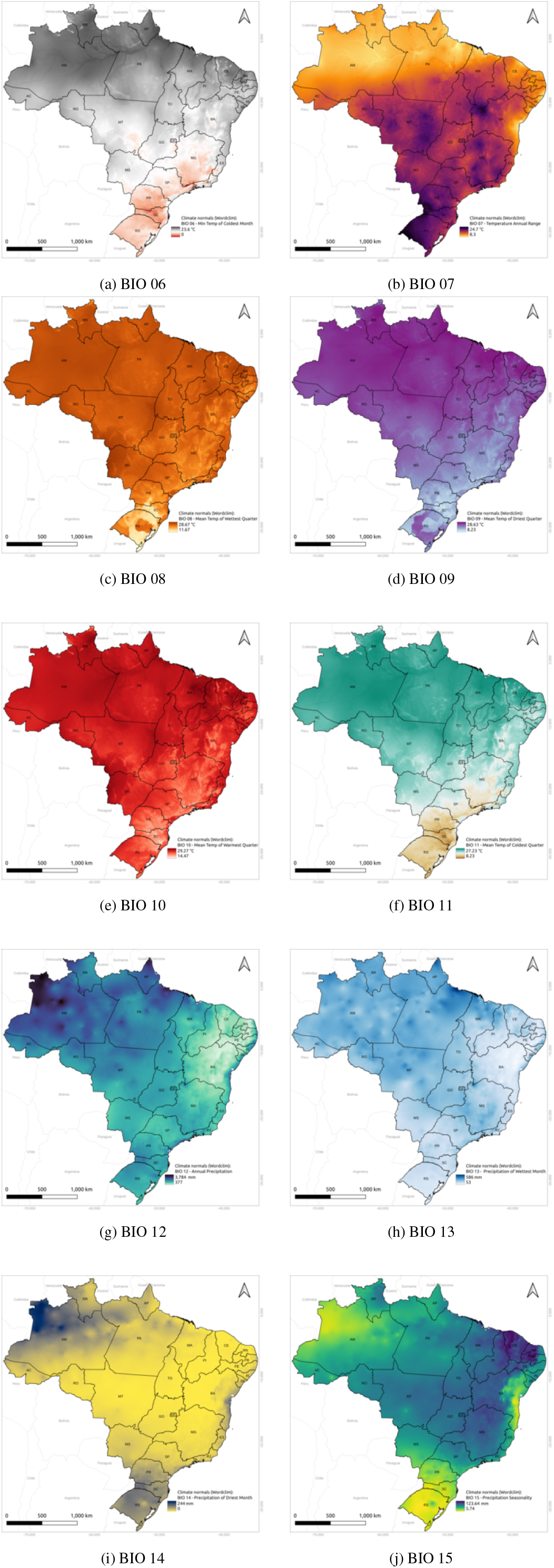
Environmental layers (II)

**Figure A.14:**
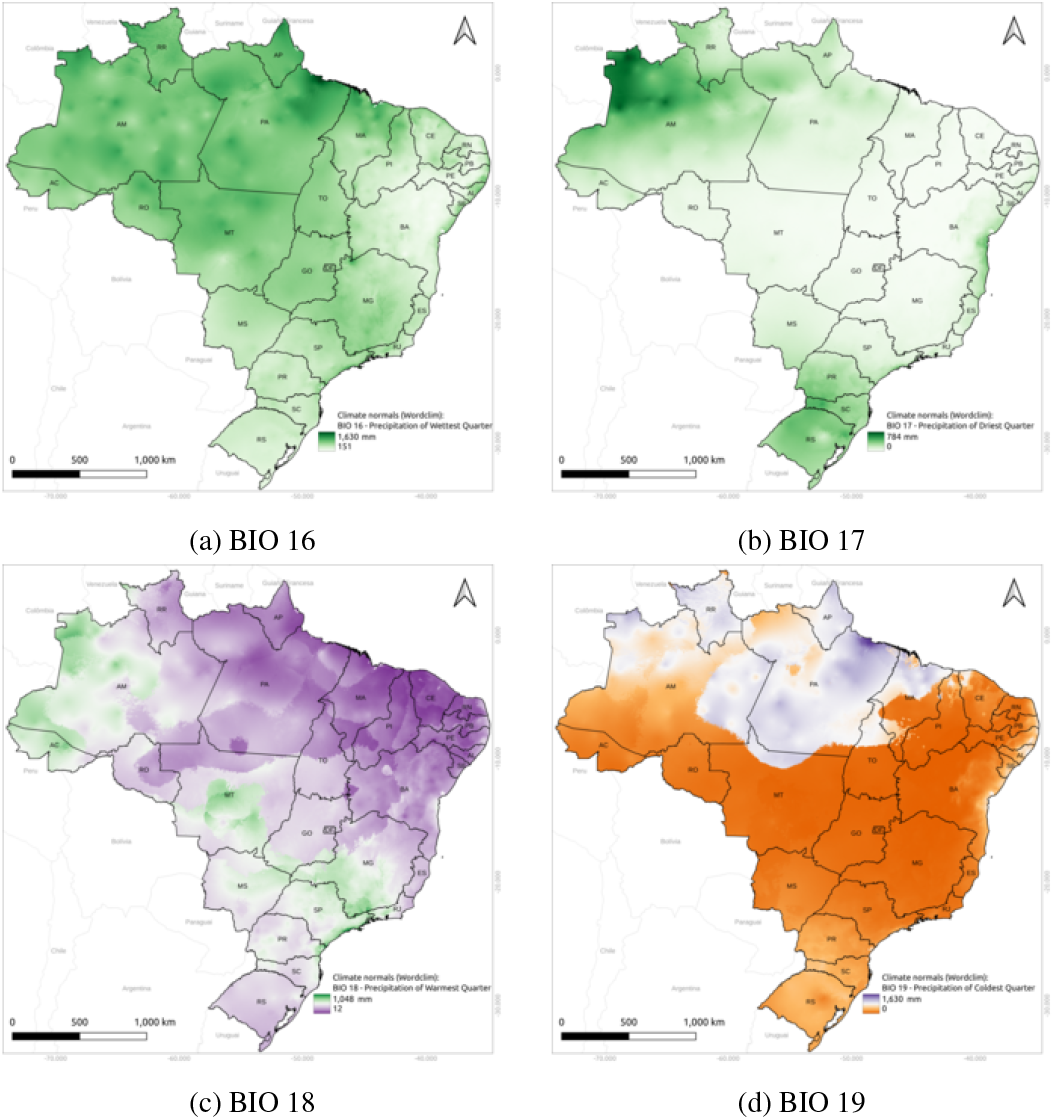
Environmental layers (III)

